# Which COVID policies are most effective? A Bayesian analysis of COVID-19 by jurisdiction

**DOI:** 10.1101/2020.12.01.20241695

**Authors:** Phebo Wibbens, Wesley Wu-Yi Koo, Anita M. McGahan

## Abstract

This paper reports the results of a Bayesian analysis on large-scale empirical data to assess the effectiveness of eleven types of COVID-control policies that have been implemented at various levels of intensity in 40 countries and U.S. states since the onset of the pandemic. The analysis estimates the marginal impact of each type and level of policy as implemented in concert with other policies. The purpose is to provide policymakers and the general public with an estimate of the relative effectiveness of various COVID-control strategies. We find that a set of widely implemented core policies reduces the spread of virus but not by enough to contain the pandemic except in a few highly compliant jurisdictions. The core policies include the cancellation of public events, restriction of gatherings to fewer than 100 people, recommendation to stay at home, recommended restrictions on internal movement, implementation of a partial international travel ban, and coordination of information campaigns. For the median jurisdiction, these policies reduce growth rate in new infections from an estimated 270% per week to approximately 49% per week, but this impact is insufficient to prevent eventual transmission throughout the population because containment occurs only when a jurisdiction reduces growth in COVID infection to below zero. Most jurisdictions must also implement additional policies, each of which has the potential to reduce weekly COVID growth rate by 10 percentage points or more. The slate of these additional high-impact policies includes targeted or full workplace closings for all but essential workers, stay-at-home requirements, and targeted school closures.

## Main Text

Many countries, regions, and U.S. states (called ‘jurisdictions’ here) have implemented COVID-control policies such as stay-at-home requirements, workplace closings, school closings, restrictions on gatherings, international travel controls, testing and contact tracing. In this paper, we report on the results of a Bayesian analysis on which policies are most important to infection control. The model generates estimates of the marginal impact of each policy in a jurisdiction after accounting for (i) the overall portfolio of policies adopted by the jurisdiction, (ii) the levels at which the policies are implemented, (iii) the rigorousness of compliance within the jurisdiction, (iv) the jurisdiction’s COVID infections, COVID deaths, and excess deaths, and (v) the performance of the portfolio of policies in other jurisdictions. The purpose is to inform decisions about changing both the level and type of policies.

We find that, across the 40 jurisdictions, the relevance of each of the policies – and the levels at which they must be implemented to drive COVID infection growth to below zero – depends sensitively on compliance in the general population with a core group of policies that are relatively less constraining of social interaction than others. We call these socially tolerable policies the “core” group in our analysis. These include the cancellation of public events, the restriction of gatherings to fewer than 100 people, recommendation to stay at home, recommended restrictions on internal movement, implementation of a partial international travel ban, and coordinated public information campaigns. However, these relatively tolerable core policies are insufficient alone for COVID control in almost all jurisdictions. At least several of the higher-impact, harder-to-tolerate, more restrictive policies must be implemented to drive growth in COVID infections below zero. We refer to these additional policies, each of which must be implemented on top of the core policies to have the estimated effect, as the “additional high-impact” group. We find that, across the 40 jurisdictions, the portfolio of additional high-impact policies includes targeted or full workplace closings for all but essential workers, stay-at-home requirements, and targeted school closings. Each is estimated to reduce the virus growth rate by 10 percentage points or more. The optimal portfolio of additional high-impact policies appropriate for each jurisdiction depends sensitively on compliance with the core policies and on interactions between policies. All data, code, and modeling information used for this project are publicly available through links described below.

Assessing the consequences of changing policies depends on accurate information about infections and deaths [1,2]. Barriers to obtaining information about infections have been extensive, and include insufficient kit supply, test insensitivity, early underestimation of the importance of testing [3], and virus-shedding by asymptomatic patients who do not present themselves clinically [4]. Deaths from COVID have also been underreported as a result of non-diagnosis, non-treatment, and structural barriers. The model on which our assessments are based (described in the Model and Data section and the Supporting Information) involves Bayesian inference about historical levels of infection inferred from both COVID deaths and excess deaths occurring a few weeks subsequently. The model is tuned through simulations to achieve goodness-of-fit by jurisdiction. The model uses global information on case fatality rates [5] and infection-to-death lags that are nested probabilistically in models of transmission within jurisdictions. Data on weekly COVID infections and deaths by country and U.S. state is drawn from the Johns Hopkins University COVID dashboard. Data on weekly expected deaths is imported from The Economist dataset.

We incorporate the weekly levels reported in the Oxford COVID-19 Government Response Tracker (OxCGRT) database in eleven categories: School closing, workplace closing, cancel public events, restrictions on gatherings, close public transport, stay-at-home requirements, restrictions on internal movement, international travel controls, public information campaigns, testing, and contact tracing.^1^ The level of each policy is assessed daily on a scale that can include up to 4 stages. The lowest level of restriction is level 1 and the greatest level is level 4. The levels are customized for each policy. For example, level 1 of international travel controls is screening arrivals whereas level 4 is closing borders to international travel.

The first set of results is a report on the average impact on COVID infection of each of the eleven policies across all the represented jurisdictions in our model. The results depend on the simultaneous deployment of the various policies across jurisdictions. The second result is a report on the percentage of jurisdictions over time with a given policy in place. The third result is the current status of new infections and growth in infections by jurisdiction. Fourth, we estimate the weekly base growth rate in infections for each jurisdiction under the hypothetical scenario that each adopts four basic policies at level 1, i.e., school closing, workplace closing, cancel public events, and restrictions on gatherings. This allows us to estimate the base rate of compliance with the portfolio of policies by jurisdiction, which in turn enables estimation of the relative impact of each policy for relatively more vs. less compliant jurisdictions. Fifth, we estimate the extent to which a set of widely adopted core policies is effective in locally containing the pandemic, and which additional high-impact policies might potentially be needed to do so. The Supporting Information, which details the model, also reports the weekly new infections and deaths for each jurisdiction as well as additional sensitivity and robustness checks.

## Existing Research

Three streams of existing research are most relevant to our study: modeling studies that estimate and predict infection and death rates, medical studies that investigate the important drivers of disease spread, and studies that estimate the effectiveness of specific policies.

### Epidemiological models

Many modeling approaches have been adopted to estimate infection rates, death rates, and disease spread. A well-established class of models relies on differential equations to analyze disease dynamics within susceptible, exposed, infected, and recovered (SEIR) “compartments” of populations [6–11]. The SEIR model and its variants (e.g., the SIR and SEIRS models) assume that the flow rates between compartments in the population are reflected in the history of the disease [9,10]. The SEIR framework has been used to study past COVID transmission and forecast future evolution [6,7,11,12]. Its variants have also produced insight on the importance of accounting for presymptomatic and asymptomatic individuals in order to curb COVID spread [13,14]. Challenges associated with SEIR-type models are that it is difficult to calibrate models (especially at initial stages when data is lacking), to predict a future trajectory of pandemic growth, and to predict policy impacts due to uncertainty in low-activity populations [15].

Despite the limitations, SEIR-type models can serve as a basis for Bayesian inference. For example, Dehning and colleagues combined Bayesian methodology with an SIR model, which is formulated in terms of coupled ordinary differential equations and enables fast evaluation [16]. Similarly, Ray and colleagues applied a Bayesian extension of the SEIR model, eSIR, to forecast future case counts and study the relative impact of lockdown duration on infections [17]. In the implementation of their model, Markov chain Monte Carlo (MCMC) simulations of the Bayesian model provides a posterior estimation of parameters and prevalence of the compartments in the SIR model. Another approach involves spatial modelling such as in the Global Epidemic and Mobility Model (GLEAM) [18–20]. This approach uses a network-based methodology and assumes that the global population can be divided into sub-populations situated around major hubs (e.g., airports and train stations). Within each sub-population, these models apply a framework similar to SEIR models through compartmental representation of the disease.

### Medical studies that are relevant to policies

A second stream of related research is comprised of medical studies that explore how the virus is transmitted. In contrast to modeling studies that customarily make use of population-level data, these medical studies pinpoint person-to-person transmission mechanisms through clinical verification within identifiable groups of individuals and/or households. An extensive amount of research has been conducted to describe and measure the airborne transmission of COVID [21–24]. In particular, researchers have highlighted the difference between indoor transmission and outdoor transmission, with the former as a predominant driver of disease transmission and the latter as a possible transmission channel [25–27]. Research has also highlighted the significant proportion of virus carriers who are asymptomatic [4,28,29]. Additionally, researchers have investigated disease transmission among different population segments. For instance, research shows that the infection rate is generally lower for children than for adults, and that children tend to catch the virus from adults and exhibit a similar level of symptoms as adults rather than disproportionately serving as asymptomatic carriers [30–33]. The Bayesian model on which we rely is designed to account for these findings.

### Policy studies

A number of studies have investigated the impact of health and non-health policies on COVID spread. This stream of research typically combines models with actual infection and death data to gauge policy effectiveness. For instance, Badr and colleagues find that reduced mobility was associated with reduced COVID growth rates in 20 U.S. counties [34]. Similarly, Bonardi and colleagues study lockdown effects on disease spread and found that reducing movement within countries was an effective control method for developed economies [35]. Meanwhile, other studies point to the lack of effectiveness of certain policies. For instance, Gostic and colleagues find that traveler screening was not particularly effective at containing the pandemic due to a large number of presymptomatic and asymptomatic travelers [36]. Facemask policies are also unlikely to help a country reach required thresholds of disease containment if the policies are loosely enforced and mask wearing remains a voluntary practice [37].

Another topic that has received attention is the impact of school closures on COVID spread. Studies have found mixed results. Some have shown that closing schools in U.S. states led to lower case number and infection rates [38]. Others have shown that, in Canada and Australia, school closures may have limited impact on reducing disease spread [39,40]. A review paper systematically examines research on the effectiveness of school closures at containing coronavirus outbreaks and concludes that school closures do not contribute to the control of the pandemic, especially in comparison to policies that broadly restrict the movement of adults [41].

While most policy studies focus on a specific set of policies or on a particular country, some studies are broader in scope. For example, Hsiang and colleagues investigate policies (non-pharmaceutical interventions) across six countries using econometric methods and conclude that different types of policies, when combined together, led to significantly reduced growth in infections [42]. Flaxman and colleagues study European countries and find that current policy interventions drive the COVID reproduction number below one [43]. Finally, Warne and colleagues have conducted one of the broadest COVID policy studies in existence: they use a stochastic model to examine COVID spread in 158 countries and associate delayed policy responses with rising cases [44].

## Methods and Data

Prior policy papers generally analyze the effectiveness of COVID control within a geographic area rather than assess how individual policies contribute across jurisdictions to the achievement of COVID control. This study is among the first to study how types of policies – implemented with varying intensity levels – perform across a large number of jurisdictions. In the sections below, we describe the model, data, and distinctive features of the approach.

### Epidemiology

The model is based on an exponential growth curve of infection rates by jurisdiction. This curve arises from a parsimonious SIR model in which the susceptible population (S) is assumed to be substantially greater than the infected (I) or recovered (R) population. The model assumes that, in most jurisdictions, just a minority of the population has been infected, and that herd immunity (i.e., more than 50% of the population has been infected) is remote in all jurisdictions. Under these assumptions, the growth in number of infections *I* is proportional to itself (d*I*/d*t* = *g I*), which gives rise to estimated exponential growth of the number of new infections in each period *X*(*t*) = Δ*I*(*t*) = e^*gt*^. In this equation, the variable *g* represents the growth rate of new infections and is closely related to the reproduction rate *R*. If *R* < 1, then *g* < 0, and the epidemic recedes locally. For *R* > 1 and *g* > 0, the number of infections grows exponentially. COVID control requires policies that yield a *g* below zero.

The reasons we model *g* rather than *R* are twofold. First, *g* is more informative of public-policy choices because it directly relates to key policy concerns, such as how many new infections and deaths must be expected, or how long a certain policy needs to be in place to reduce the number of new infections to a given threshold. Second, *g* is more readily observable from the data. Estimating *R* requires making additional assumptions about the time lag between original infection and re-infections by a given individual that are not relevant for policy analysis.

We assume that the growth rate for each jurisdiction *i* in week *t* is:

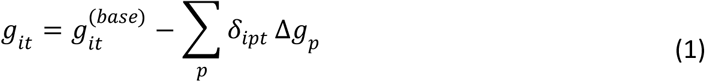

The first term represents the base growth rate by jurisdiction, while the second term represents the growth rate reduction arising from the implemented policies. The base growth rate is represented by the equation *g*_*it*_(base) = *g*_*i*_^(0/1)^ + Δ*g*_*it*_(id), which consists of two parts. The term *g*_*i*_(0) represents COVID growth prior to the implementation of any policies (e.g., equivalently to *R*_0_ in SIR models). After jurisdictions have implemented basic containment policies, *g*_*i*_(0) is replaced by *g*_*i*_(1) (see below for detail on policy coding). The second part, Δ*g*_*it*_^(id)^, represents the idiosyncratic growth rate. This term captures any jurisdiction-specific changes over time in COVID growth that cannot be attributed to the policies coded in our database. For this term, we assume a second-order autoregressive process (AR(2)). Mathematical details are provided in the Supporting Information.

The term ∑_*p*_ *δ*_*ipt*_ Δ*g*_*p*_ in Equation (1) represents the effect of further containment policies as coded in the OxCGRT database. The variable *δ*_*ipt*_ = 1 if jurisdiction *i* had policy *p* in place in week *t*, and *δ*_*ipt*_= 0 otherwise. Δ*g*_*p*_ ≥ 0 is the estimate for the average effect of each policy.

The growth rate is determined by the weekly change in new infections in logarithms: *g*_*it*_ = log(*X*_*it*_) – log(*X*_*i,t*-1_). Because the number of infections is not directly observable, it is inferred from three different observations by jurisdiction-week: the number of reported COVID cases, the number of reported COVID deaths, and the number of excess deaths. The model considers that reported COVID cases and deaths may be significantly understated, and that the level of underreporting may evolve with changing testing policies. The model also accounts for the lag between infections on the one hand and reported cases and deaths on the other.

### Data

We use two sets of input data: policy data to code the variable *δ*_*ipt*_ and epidemiological data to infer the level of new infections *X*_*it*_. The policy data are based on the OxCGRT database [45], which codes multiple levels of policy responses across many categories for all countries in the world and all U.S. states. The database is publicly available (see links provided at the end of this paper).

Table 1 lists the coding of the levels in the OxCGRT database (the information in this table is copied directly from the Oxford team’s website on GitHub) of policies that related to virus containment. For school closures (category C1), we adapted the coding to include the summer holidays for countries in the northern hemisphere, with level 3 representing closure in all geographies and level 2 in some (because of regional spread of vacations within a jurisdiction). The highest level prevails between the original OxCGRT and the holiday coding (e.g., if OxCGRT codes level 3 for school closures, while holidays are level 2, then we keep the original level 3 coding). Figure 1 displays the coding of weekly policy by jurisdiction in the model.

**Table 1.** Policy coding used in the OxCGRT database. (continued on next page) Adapted from: https://github.com/OxCGRT/covid-policy-tracker/blob/master/documentation/codebook.md, retrieved on 23 September 2020.

| ID | Description | Policy level |
| --- | --- | --- |
| C1 | Record closings of schools and universities | 0 - no measures<br>1 - recommend closing<br>2 - require closing (only some levels or categories, eg just high school, or just public schools)<br>3 - require closing all levels<br>Blank - no data |
| C2 | Record closings of workplaces | 0 - no measures<br>1 - recommend closing (or recommend work from home)<br>2 - require closing (or work from home) for some sectors or categories of workers<br>3 - require closing (or work from home) for all-but-essential workplaces (eg grocery stores, doctors)<br>Blank - no data |
| C3 | Record cancelling public events | 0 - no measures<br>1 - recommend cancelling<br>2 - require cancelling<br>Blank - no data |
| C4 | Record limits on private gatherings | 0 - no restrictions<br>1 - restrictions on very large gatherings (the limit is above 1000 people)<br>2 - restrictions on gatherings between 101-1000 people<br>3 - restrictions on gatherings between 11-100 people<br>4 - restrictions on gatherings of 10 people or less<br>Blank - no data |
| C5 | Record closing of public transport | 0 - no measures<br>1 - recommend closing (or significantly reduce volume/route/means of transport available)<br>2 - require closing (or prohibit most citizens from using it)<br>Blank - no data |
| C6 | Record orders to "shelter-in-place" and otherwise confine to the home | 0 - no measures<br>1 - recommend not leaving house<br>2 - require not leaving house with exceptions for daily exercise, grocery shopping, and 'essential' trips<br>3 - require not leaving house with minimal exceptions (eg allowed to leave once a week, or only one person can leave at a time, etc)<br>Blank - no data |

**Table 1.** (continued from previous page)
| ID | Description | Policy level |
| --- | --- | --- |
| C7 | Record restrictions on internal movement between cities/regions | 0 - no measures<br>1 - recommend not to travel between regions/cities<br>2 - internal movement restrictions in place<br>Blank - no data |
| C8 | Record restrictions on international travel | 0 - no restrictions<br>1 - screening arrivals<br>2 - quarantine arrivals from some or all regions<br>3 - ban arrivals from some regions<br>4 - ban on all regions or total border closure<br>Blank - no data |
| H1 | Record presence of public info campaigns | 0 - no COVID-19 public information campaign<br>1 - public officials urging caution about COVID-19<br>2- coordinated public information campaign (eg across traditional and social media)<br>Blank - no data |
| H2 | Record government policy on who has access to testing | 0 - no testing policy<br>1 - only those who both (a) have symptoms AND (b) meet specific criteria (eg key workers, admitted to hospital, came into contact with a known case, returned from overseas)<br>2 - testing of anyone showing COVID-19 symptoms<br>3 - open public testing (eg "drive through" testing available to asymptomatic people)<br>Blank - no data |
| H3 | Record government policy on contact tracing after a positive diagnosis | 0 - no contact tracing<br>1 - limited contact tracing; not done for all cases<br>2 - comprehensive contact tracing; done for all identified cases |

**Figure 1.**
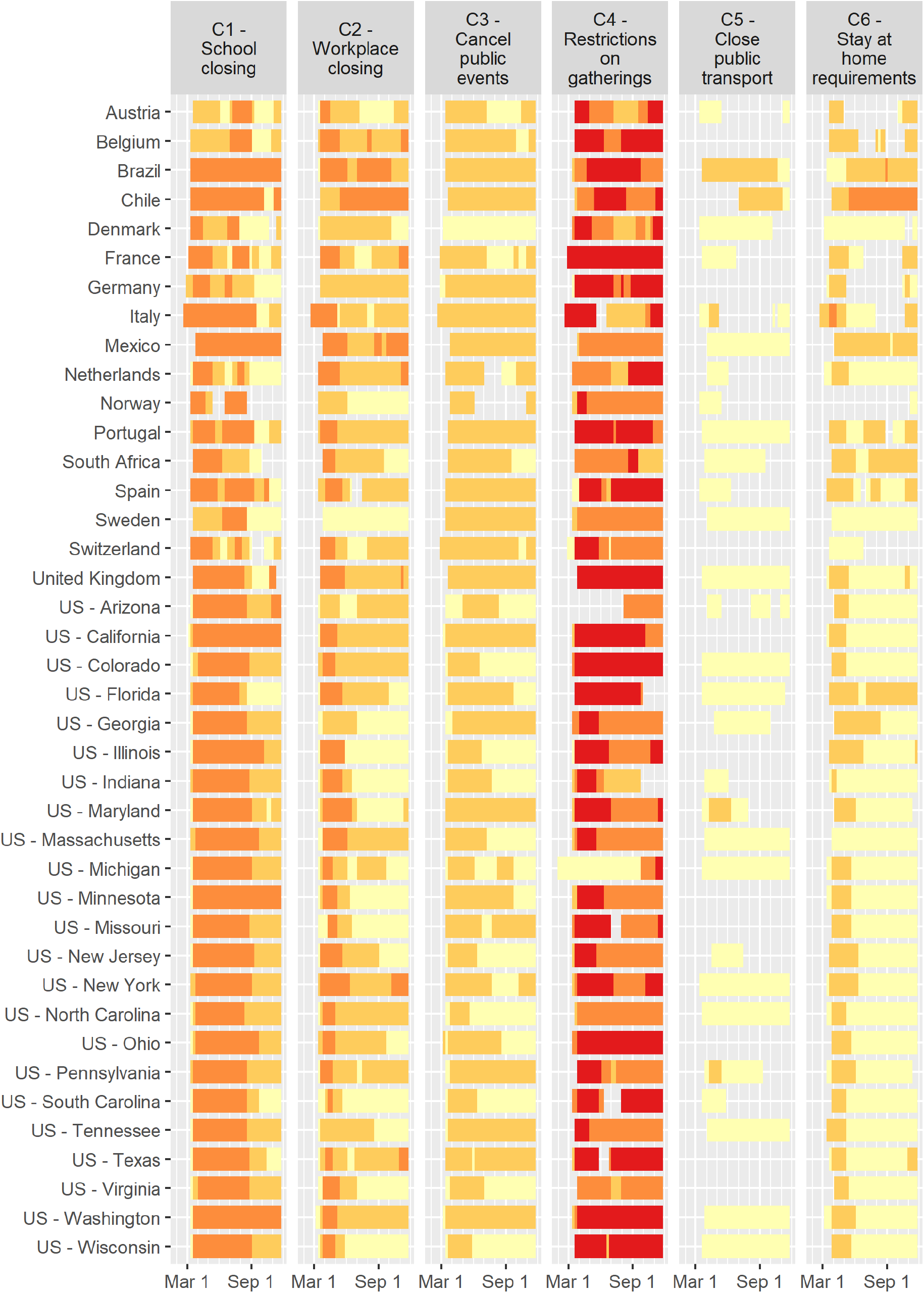
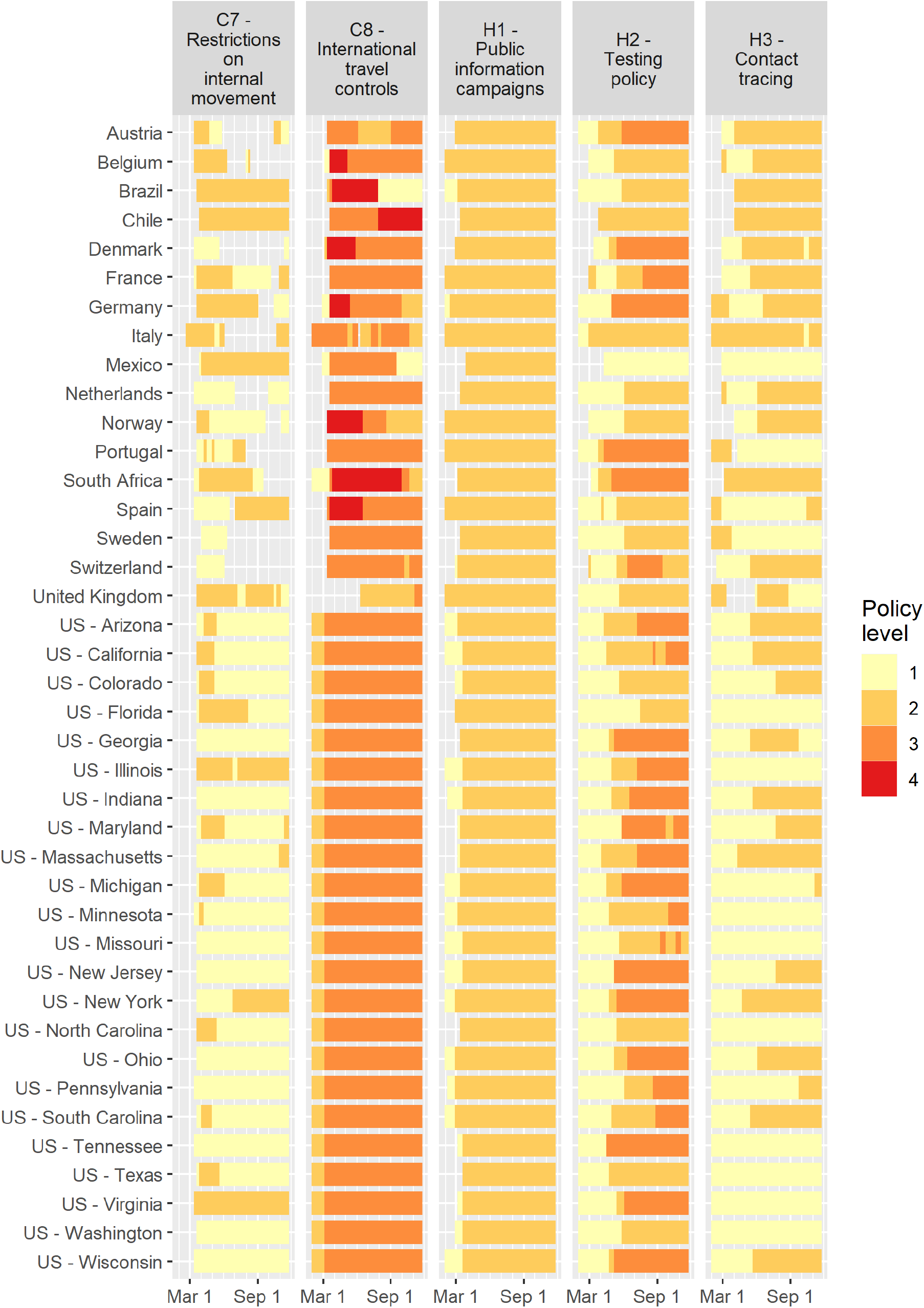
COVID policy levels by week based on OxCGRT database.

The model treats the levels cumulatively. For instance, if a jurisdiction is coded as level 2 on policy H2 (virus testing) in a certain week, then we code *δ*_*ipt*_ = 1 for both level 1 and level 2, and *δ*_*ipt*_ = 0 for level 3. In other words, the effect of H2 policies on the overall growth rate *g*_*it*_ is modelled as Δ*g*_*p=*H2-1_ + Δ*g*_*p=*H2-2,_with H2-*n* representing policy level *n* for category H2.

Because significant policy efforts in essentially all jurisdictions began with C1 (school closing), C2 (Workplace closing), C3 (Cancel public events), and C4 (restrictions on gatherings), we use the implementation of any of these policies at level 1 or above as the criterion for shifting the time-constant part of the base growth rate from *g*_*i*_^(0)^ (free spread of the virus corresponding to *R*_0_) to *g*_*i*_^(1)^ (spread of the virus with only basic policies in place). Because C1-C4 are implemented almost simultaneously at an initiating level 1 or greater, they are highly collinear in the estimation of Equation <u>(1)</u>—each has a correlation coefficient of 0.85 or higher with other policies. Therefore, we exclude these policies in the estimation of Δ*g*_*p*_. For the same reason we also exclude policy levels that are implemented in fewer than 5% of all policy-weeks.^2^ This results in 23 distinct policy levels that we include in our analysis.

The epidemiological data arise from two sources. COVID case and death data are taken from the Johns Hopkins Coronavirus Resource Center to calculate weekly counts of new cases and deaths by jurisdictions. Excess death data are drawn from The Economist, which reports weekly and monthly total deaths above what was projected based on pre-COVID trends for a limited set of countries and U.S. states. The excess deaths over expected deaths serve as a proxy for the underreporting of COVID-19 deaths.

In our analysis, we include all countries and U.S. states with at least 5 million residents for which The Economist reports excess deaths weekly. Forty jurisdictions meet these criteria. The Supporting Information contains robustness checks on the criteria for including jurisdictions.

### Estimation

Both policy and epidemiological data are used to estimate the effect of various policies on COVID control. The primary variable of interest is Δ*g*_*p*_, which represents the marginal effect of each implemented policy, *p*, on reducing the weekly growth rate of infections. We also report results for *g*_*it*_^(base)^, the base growth rate in infections independently of the implemented policies coded in Δ*g*_*p*_. Finally, we use the model to make inferences by jurisdiction of weekly new infections *X*_*it*_ and the infection growth rate *g*_*it*_.

The Bayesian model generates point estimates (posterior median) and 95% intervals (posterior quantiles) for each of the variables of interest. Equation (1) is linear, and thus would be estimable through ordinary least squares (OLS) regression, but Bayesian estimation confers several advantages. First, the dependent variable (DV) in Equation (1), *g*_*it*_ = log(*X*_*it*_) – log(*X*_*i,t*-1_), is not directly observable because we do not know the actual level of new infections *X*_*it*_. Therefore, this variable must be estimated probabilistically based on observations of reported COVID cases, reported COVID deaths, and excess deaths. Bayesian modeling is uniquely suited to accomplish this. For the reported case count, *Y*_*it*_^(case)^, we adopt a negative binomial model with rate parameter *μ* and overdispersion parameter *ϕ*:

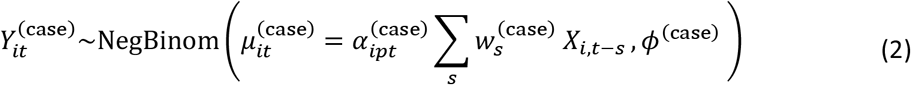

The rate parameter *μ* represents the expected rate of new cases based on the fraction of total cases *α*_*ipt*_^(case)^ that is reported in jurisdiction *i* with testing policy (H2) *p* in week *t*; the fraction of cases *w*_*s*_ that is reported with a lag of *s* weeks; and the actual number of infections *X*_*it*_. Similar negative binomial models are assumed for reported COVID deaths and excess mortality.

One main advantage of the Bayesian approach is that we can use the observed data to make stochastic inferences (with 95% intervals) about the three parameters *α, w*, and—most importantly—the number of new infections *X*, without any further assumptions. The Markov Chain Monte Carlo (MCMC) estimation essentially performs hundreds of simulations of parameter combinations, calculating the likelihood of each combination given the observed data of reported cased, reported deaths, and excess deaths.

A second advantage of the Bayesian model is that it is robust to outliers [46: p.435]. This is important because extreme outliers may influence the inference despite the feature of the negative binomial distribution to provide some protection against outliers by allowing over-dispersion as compared to the Poisson distribution. For instance, in some weeks, a few jurisdictions report very few or no cases due to bureaucratical constraints on disclosure. In the Bayesian model, no distortion arises as a result of this practice through the assumption that an observation is randomly generated with a small probability *p*_outlier_, and that the observation is drawn from the distribution represented by Equation <u>(2).</u> The model assumes *p*_outlier_ = 10^−3^ for reported COVID cases and deaths (no assumption is necessary for excess deaths because these do not exhibit outliers). In the Supporting Information we also present the results of robustness checks using outlier probabilities of 10^−2^ and 10^−4^.

Third and finally, Bayesian analysis accommodates further constraints and/or stochastic assumptions. For instance, we impose a constraint that policy effects reduce rather than increase infection by assuming Δ*g*_*p*_ ≥ 0. Other constraints include that the fraction *α* of cases/deaths that is reported and the lag *w* take on values between 0 and 1, and that the sum of lag weights *w* equals 1. Similarly, the estimation accommodates reliance on the AR(2) process for the idiosyncratic term Δ*g*_*it*_^(id)^ (see Equation (S3) of the Supporting Information for further details).

## Results, Discussion, and Implications for Policy

For the population of countries and U.S. states included in our analysis, the marginal impact Δ*g*_*p*_ of each policy *p* is represented in Figure 2. Each row of the figure represents one of the eleven types of policies reported in the OxCGRT database. The bars represent the median estimate of each policy’s impact on reduction in the weekly average growth rate in infections. Each bar is colored to represent the levels of intensity with which the policies were implemented (see Table 1 for descriptions of the levels). The lines represent the 95% interval around each estimate at the maximum policy level. Overall, Figure 2 shows which of the eleven policies – and the level of intensity of those policies – produces the greatest relative impact on reducing growth in COVID infections across the jurisdictions that we study.

**Figure 2.**
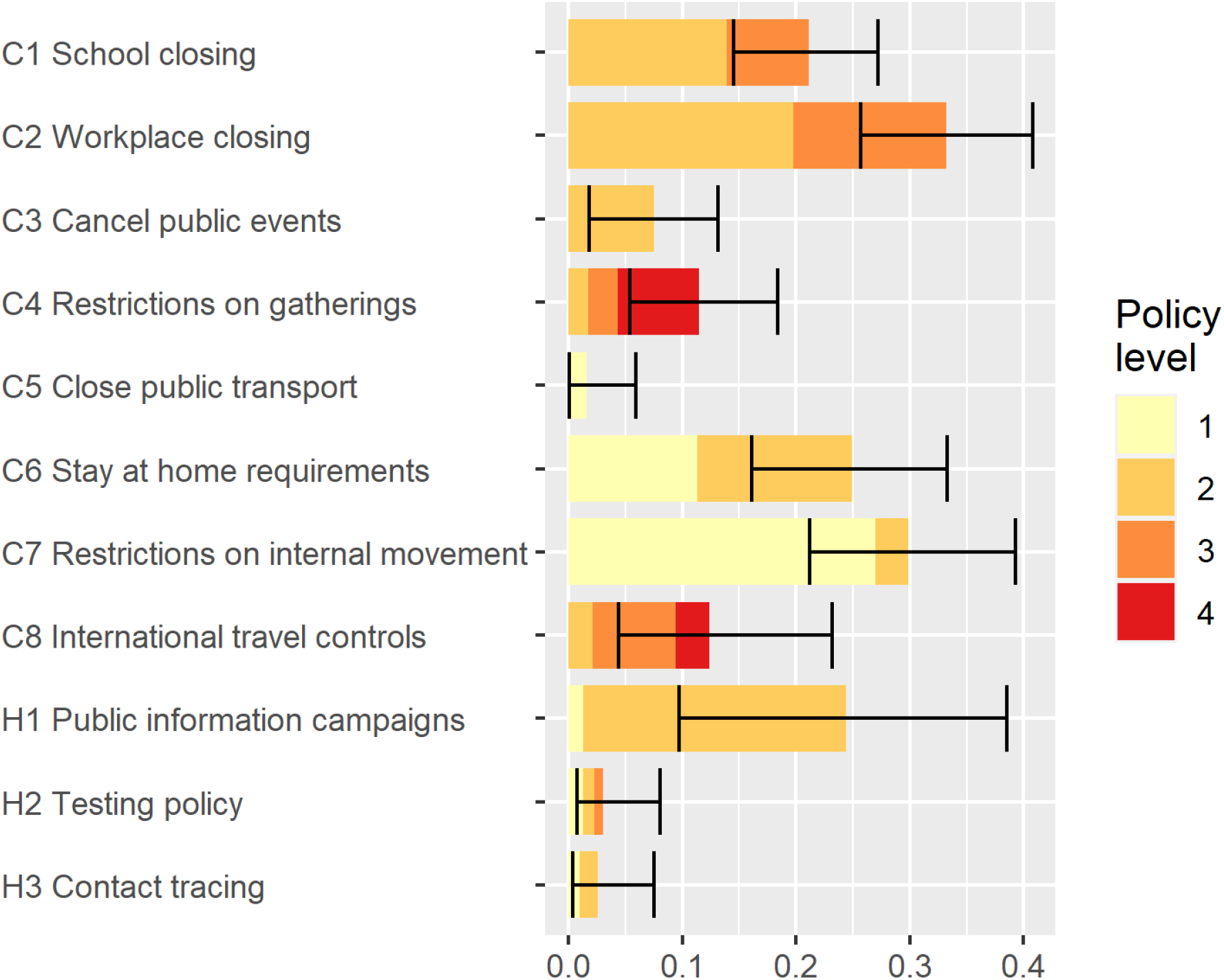
Marginal effect sizes of policy measures on reducing weekly growth rates. Bars = median estimate by policy level; Lines = 95% intervals for maximum policy level

Several important results are evident in Figure 2. First, five of the policies have relatively large impact when fully implemented at their highest levels: Workplace closing, restrictions on internal movement, stay-at-home requirements, public information campaigns, and school closings. Four of the five are associated with lower growth of infections even at relatively low levels, but public information campaigns (H1) are most impactful at the highest recorded level (which in this case is level 2, i.e., coordination across traditional and social media). This policy was implemented early in the pandemic by all jurisdictions in our analysis and is part of the core group (see Figure 1). Second, constraints on the movements of adults through C2 (workplace closings for all but essential workers), C7 (restrictions on internal movements), and C6 (stay-at-home requirements with exceptions for daily exercise, grocery shopping, and ‘essential’ trips) have the greatest marginal impact at intermediate levels. These results are consistent with prior results from the literature suggesting that control of adult-to-adult transmission is central to COVID remediation. Third, the largest marginal effect of school closing (C1) is at level 2 (closing of some categories, such as universities and/or high schools) while a level 3 policy (closing of all schools) provides and additional but lower marginal benefit. Fourth, testing policy and contact tracing are not strongly associated with COVID controlling for the 40 jurisdictions that we study.

Figure 3 represents the current status at the time of writing (November 22, 2020) of the results of the implemented policies. Several jurisdictions have elevated numbers of new infections *X*_*it*_. For Italy and Switzerland, new infections are more than 200 per 10,000 people weekly, which is close to the highest numbers of infections in the early stages of the pandemic in Spain, New York, and New Jersey. Growth rates of new infections are highest in Sweden, Colorado, and Pennsylvania. In these jurisdictions, new infections are growing at a weekly rate around or above *g*_*it*_ = 0.4, which corresponds to an increase of 49% new cases every week (e^0.4^ – 1 = 0.49). In other jurisdictions, the growth rate is closer to or below zero (although presently the rate is estimated to be below zero with at least 95% certainty only in Belgium, Chile, Denmark, and France). These differences arise from variation in the policies in place as represented in Figures 1 and 2 as well as compliance, which is driven in part by jurisdiction-specific factors such as population density.

**Figure 3.**
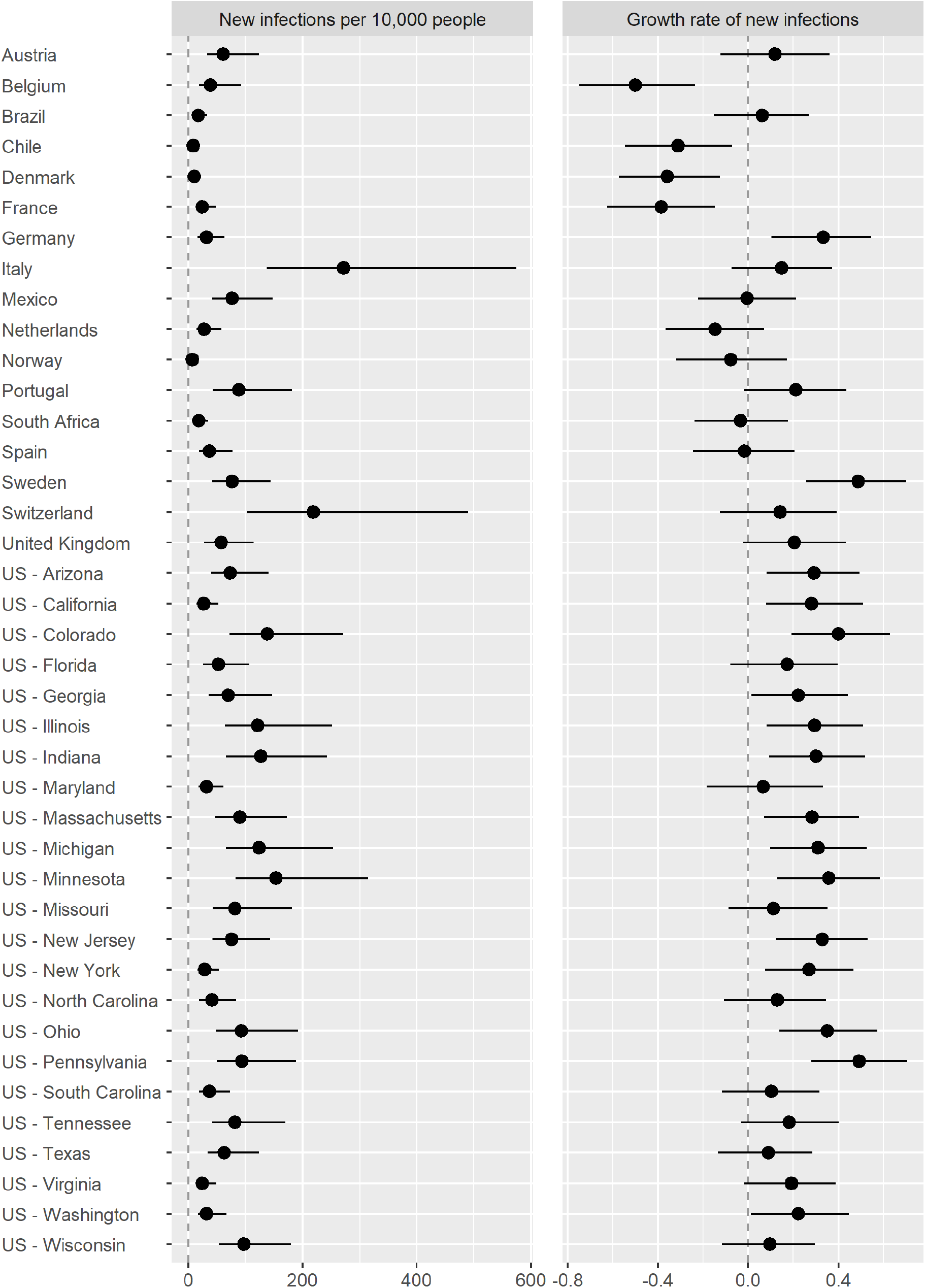
Weekly rate of new infections and their growth by jurisdiction as of November 22, 2020. Dots = median estimates; Lines = 95% intervals

Figure 4 provides insight into compliance. The figure reports on estimates of the base growth rate *g*_*it*_^*>*(base)^ by jurisdiction in the hypothetical situation in which only level 1 policies were implemented on school closing, workplace closing, canceling public events, and restrictions on gatherings (see Equa<u>tion (1)</u>). Especially earlier in the pandemic, Sweden stood out as a country in which these base policies appeared to have had a great impact on lowering growth in infections, resulting in a very low base growth rate *g*_*i*_^(1)^. The model does not, however, offer reasons for this outcome. For example, Sweden’s relatively low population density, weather characteristics, and demographics likely play a role in the result. In recent months, Sweden’s base rate of growth in infection is estimated to have increased, while other jurisdictions such as Denmark and South Africa have estimated *g*_*it*_^(base)^ that have declined to levels below that of Sweden. By contrast, Brazil, Chile, Mexico, and Spain have had at least some periods in which implementation of only base policies would likely have not had as strong an impact on COVID control.

**Figure 4.**
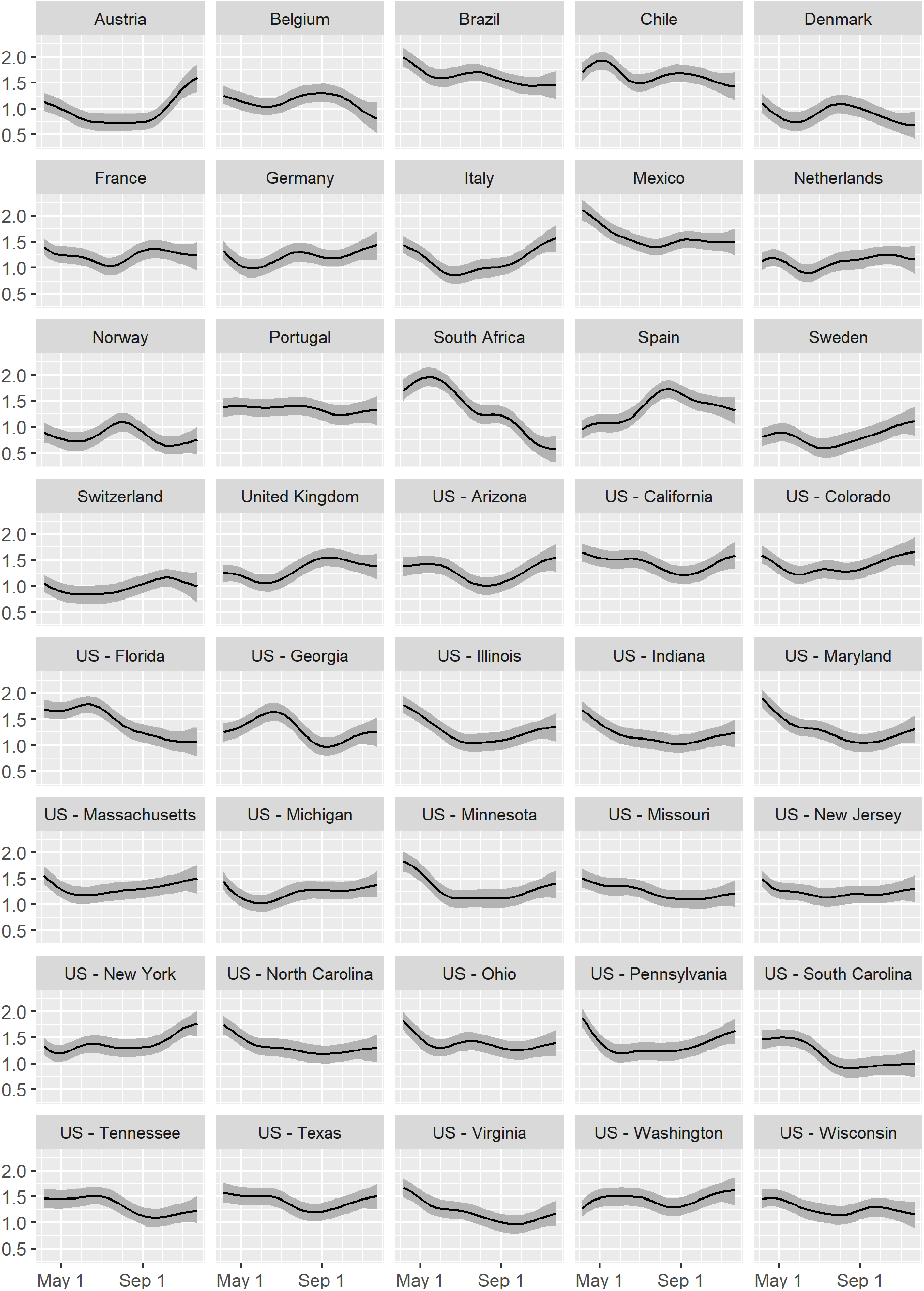
Growth rate *g*^(base)^ after jurisdictions have started to implement one of the base policies (level 1 of C1-C4). A lower number indicates a higher effectiveness to contain the virus. The total growth rate *g*_*it*_ of the virus is the number in this chart minus the sum of the effects of implemented policies in Figure 2. Lines = median estimates; Grey bands = 95% intervals

Figure 5 illustrates the implications of compliance in a jurisdiction for the additional high-impact policies that must be implemented above the core group to reduce COVID growth below zero. To read the figure, please begin by examining the *g*^(base)^ for each category of jurisdiction: the most compliant 10%; the median by compliance; and the least compliant 10%. For jurisdictions in each of these categories, the figure shows how much reduction in the growth rate *g* has been accomplished on average through implementation of the core policies, which have been widely adopted across jurisdictions because of their social tolerability. These core policies include cancellation of public events (C3 – 2), restricting gatherings to fewer than 100 people (C4 – 3), recommending stay-at-home behavior (C6 – 1), recommending no travel between regions/cities (C7 – 1), implementing a partial international travel ban (C8 – 3), instituting coordinated information campaigns (H1 – 2), and COVID testing and tracing (H2 – 2 & H3 – 2), in addition to the base policies (C1-C4 level 1).

**Figure 5.**
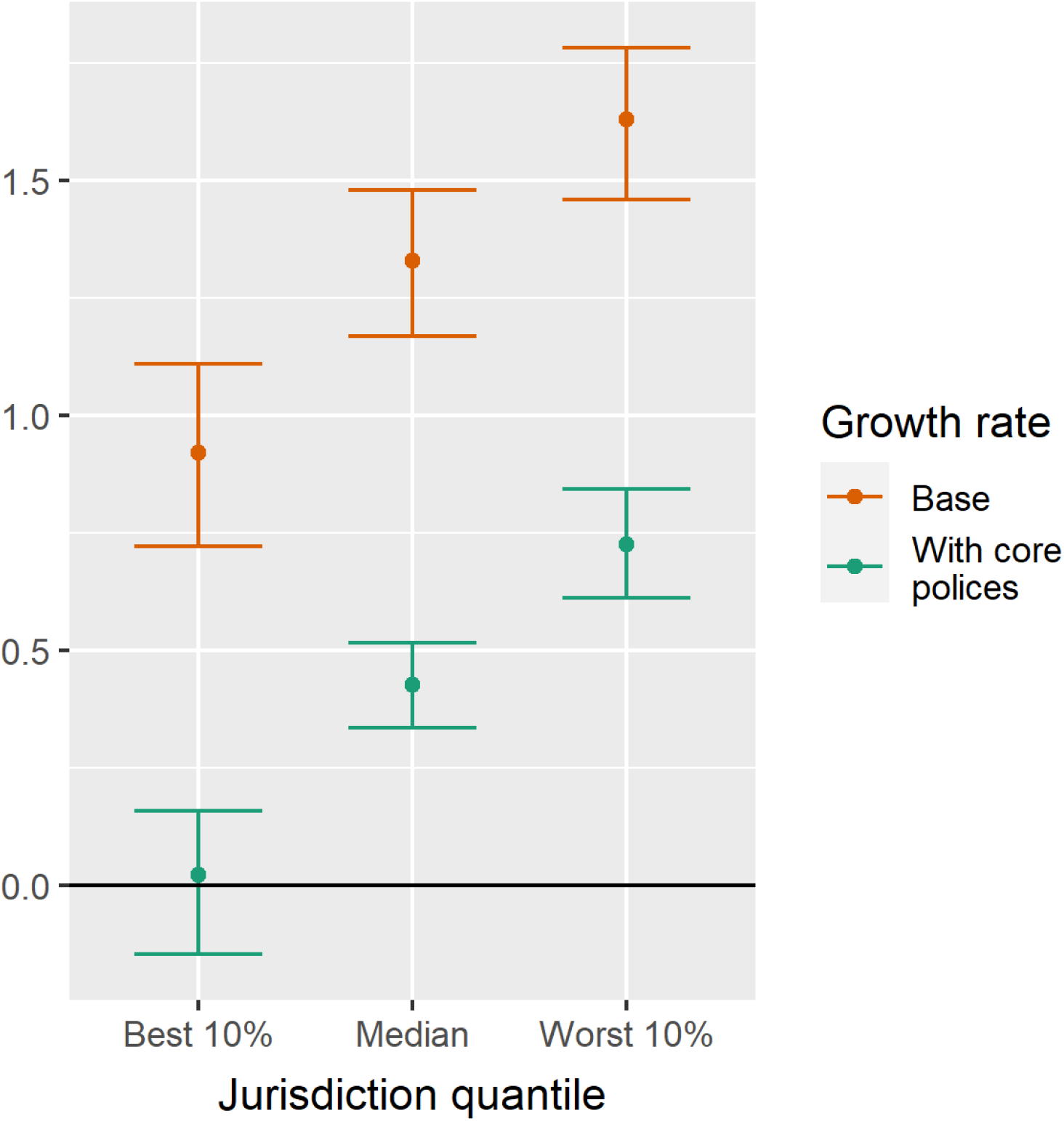
Effects of a set of core policies on weekly growth rate *g* by quantile of *g*^(base)^ among all jurisdictions. Core policies include the cumulative effects of C3 - 2, C4 - 3, C6 - 1, C7 - 1, C8 - 3, H1 - 2, H2 - 2, and H3 - 2. Dots = median estimates; Lines = 95% intervals.

The figure shows that the set of core policies lead to COVID control only for jurisdictions with strong policy compliance (i.e., the lowest *g*^(base)^). In contrast, for the median jurisdiction, the core policies are insufficient for driving COVID growth below zero despite having a strong effect, i.e., they reduce growth from an estimated 1.3 cases per week corresponding to an increase in virus spread by e^1.3^ – 1 = 270% to an estimated 0.4 cases per week corresponding to a growth of e^0.4^ – 1 = 49%. Hence, for 90% of jurisdictions, the core policies are insufficient to achieve COVID control. Almost all jurisdictions must implement more stringent policies to achieve a further reduction of the weekly growth rate *g* to below zero. Based on the marginal effects in Figure 2, the additional high-impact policies (with median estimated effectiveness Δ*g*_*p*_ above 0.1) are:

- Targeted workplace closings (C2 – 2; Δ*g*_*p*_ = 0.20)
- Full workplace closings (C2 – 3; Δ*g*_*p*_ = 0.14, on top of C2 – 2)
- Stay-at-home requirement with exceptions (C6 – 2; Δ*g*_*p*_ = 0.14)
- Targeted school closings (C1 – 2; Δ*g*_*p*_ = 0.14)

The next two most effective policies that must be implemented in addition to the core policies are (with median estimated effectiveness Δ*g*_*p*_ between 0.05 and 0.1):

- Full school closings (C1 – 3; Δ*g*_*p*_ = 0.07, on top of C1 – 2)
- Restrictions on gatherings below 10 (C4 – 4; Δ*g*_*p*_ = 0.07)

Each of these policies has significant societal costs. Unfortunately, however, unless governments can compel greater compliance with core policies, jurisdictions must implement several of these additional policies to ensure that virus spread does not grow exponentially.

As with any study, these results must be interpreted within the limitations of the methods used. One limitation is that we need to make several assumptions as described in the Methods section about disease epidemiology and the impact of government policy. In the Supporting Information, we have included several robustness checks that demonstrate that our results are relatively insensitive within reasonable parameters to specific assumptions. Second, because this is an observational study, it is inherently limited because the findings may be biased by variables that are not included in the study. For instance, the relatively limited impact of testing (H2) and contact tracing (H3) in Figure 2 could arise from the lifting of policies others than those reported in the Oxford database around the same time that H2 and H3 policies were implemented. While we strive to limit the impact of omitted-variable bias through reliance on 23 different policy levels, the findings must be interpreted with caution. Overall, the results should be interpreted with these limitations in mind and should be relied on only in conjunction with results from other studies, including computational models and controlled experiments such as those described in our review of the literature.

## Conclusion

The analysis generates several important insights. First, important differences arise across jurisdictions in infection levels, death rates, policy implementation, and compliance with policies. These differences have major implications for COVID control. The portfolio of the eleven types of policies – implemented with different levels of intensity – necessary to drive COVID growth rate below zero depends on the jurisdiction’s COVID burden and compliance, which reflects behavioral and demographic characteristics of the jurisdictions. Second, we estimate that a core set of socially tolerable policies lead to COVID control only in those jurisdictions that have unusually high levels of compliance. The socially tolerable core policies alone are meaningful and significant, but insufficient by themselves for preventing escalating growth in infections in 90% of the jurisdictions analyzed. For these jurisdictions, one or more from a set of additional high-impact but difficult-to-tolerate policies must be implemented to achieve COVID control. Third, the impact of testing and contact tracing has been lower than the impact of other policies. Fourth, for the jurisdictions covered in this analysis, the policies with the greatest marginal impact for achieving COVID control mainly involve restrictions on adults through workplace closings and stay-at-home requirements, although targeted school closings are also in the group of additional high-impact policies. Altogether, the analysis indicates that, in all but a few highly compliant jurisdictions, relatively significant social costs must be incurred to reduce COVID growth below zero. The model points to significant opportunities for cultivating deeper understanding of the drivers of compliance and of variation in the impact of policies, which are potentially rewarding topics for future research.

## Data Availability

All data is publicly available, and all code is made available on Github.

https://github.com/phebo/covid-19projections

https://github.com/OxCGRT/covid-policy-tracker

https://github.com/CSSEGISandData/COVID-19

https://github.com/TheEconomist/covid-19-excess-deaths-tracker

## Author contributions

All authors contributed equally. Wibbens conducted the modeling. Koo gathered data, did policy analysis, and wrote the literature review. McGahan was primarily responsible for the remaining writing. All authors discussed each element of the project.

## Data and materials availability

Policy data from the OxCGRT data are available at https://github.com/OxCGRT/covid-policy-tracker. Deaths and reported case data for countries was drawn from the Johns Hopkins Coronavirus Resource Center at https://github.com/CSSEGISandData/COVID-19. Data for The Economist excess death tracker are available at https://github.com/TheEconomist/covid-19-excess-deaths-tracker. The code for the model is available at https://github.com/phebo/covid-19projections.

## Footnotes

1 The Oxford data can be accessed at: https://www.bsg.ox.ac.uk/research/research-projects/coronavirus-government-response-tracker.

2 We also used several different sets of base policies for *g*_*i*_ ^(1)^ and of excluded variables. The marginal effects for these (unreported) analyses were not significantly different from those presented in Figure 2.

