## Supporting Information for "Which COVID policies are most effective? A Bayesian analysis of COVID-19 by jurisdiction"

**for**

### Bayesian methodology

The Bayesian estimation model in this paper simulates the probability distribution of a set of parameters $\theta$ based on observed data $y$ using Bayes formula $p(\theta|y) = p(\theta)\ p(y|\theta)\ /\ p(y)$. In this equation, $p(\theta|y)$, the probability distribution of the parameters given the observed data, is called the *posterior distribution*, which is the function the model simulates. $p(\theta)$ is the *prior distribution*, containing prior information about the parameters, such as earlier data on mortality. $p(y|\theta)$ is the likelihood function, which encodes how likely it is to observe certain outcomes given parameter estimates, such as the negative binomial distribution that we assume for observing a certain number of reported deaths given a level of infection rates. Finally, $p(y)$ is constant in $\theta$ and commonly disregarded in Bayesian analysis as it does not contain any further information about the inference of the parameters of interest. Further details on Bayesian estimation can be found in standard textbooks such as [1].

The detailed specification of the Bayesian model is described below. The distribution of the posterior is $p(\theta|y)$ estimated using a so-called Markov Chain Monte Carlo (MCMC) simulation. Essentially, this technique calculates the likelihood of many different parameters $\theta$ to simulate a set of draws taken from the posterior distribution. The MCMC simulation is implemented through the R-version of Stan, a state-of-the-art programming language to estimate complex Bayesian models, which uses  [2].

We estimate a total of 1000 posterior samples for each variable in $\theta$. These are drawn from four independent MCMC chains, consisting of 1000 simulations each, of which the first 500 are discarded as warm-up, and the sampled chains are thinned by 2 to conserve space. The purpose is for the model to find the variable combinations with highest likelihood. This leads to four groups of 250 independent draws from the posterior distributions. Sampling takes several hours on a high-end dedicated computer.

Table S1 shows posterior inference for several key parameters of interest. The effective sample size $n_{\text{eff}}$ and the $R_{\text{hat}}$ statistics support assessments of the proper convergence of the MCMC estimates. According to Gelman et al. [2: p.287], the effective sampling size should be at least 10 times the number of chains (40 in this case), and the $R_{\text{hat}}$ statistic close to 1, preferably below 1.1. These conditions are met for all the variables in Table S1. The role of each of these parameters in the model will be discussed further below.

Table S2 shows the variance inflation factors the policy fixed effects $\delta_{ipt}$ by policy $p$. The highest value is 5.19, which is well below the typically recommended maximum of 10. This ensures that posterior intervals are not overly inflated due to multi-collinearity of the policy implementations.

### Further details to the model

Analogous to the negative binomial distribution for reported cases (Equation (2) in the main text), we assume similar distributions for reported COVID deaths as well as total deaths:

$$Y_{it}^{(\text{death})} \sim \text{NegBinom}\left(\mu_{it}^{(\text{death})} = \alpha_{ipt}^{(\text{death})}\, m \sum_{s} w_s^{(\text{death})}\, X_{i,t-s}\,, \phi^{(\text{death})}\right) \qquad \text{(S1)}$$

$$Y_{it}^{(\text{deathTot})} \sim \text{NegBinom}\left(\mu_{it}^{(\text{deathTot})} = E_{it} + A_i + m \sum_{s} w_s^{(\text{death})}\, X_{i,t-s}\,, \phi^{(\text{deathTot})}\right) \qquad \text{(S2)}$$

In these equations, $\alpha_{ipt}^{(\text{death})}$ represents the fraction of total COVID deaths that is reported in jurisdiction $i$ with testing policy (H2) $p$ in week $t$; $m$ the mortality of a COVID infection; $w_s^{(\text{death})}$ the fraction of cases that is reported with a lag of $s$ weeks; $\phi$ the overdispersion parameters as compared to a Poisson distribution; $E_{it}$ the number of expected deaths by jurisdiction-week, and $A_i$ a time-constant adjustment factor by jurisdiction. All these parameters are estimated by the model, except for the expected deaths $E_{it}$, which is included from The Economist excess mortality database. The expected deaths are estimated based on weekly observed deaths in prior years and population developments. Full methodological details are available from the GitHub repository provided by The Economist (see weblink provided in the main text under Data and materials availability). For the mortality $m$ we assume a log-normal prior with parameters based on prior literature. We use $\mu$ = exp(0.01) and $\sigma$ = 0.5, implying a 95% interval for mortality between 0.3% and 2.7%, in line with estimates by Wilson et al. [5, main text].

For all other parameters we use uninformative priors. This ensures that the posterior distributions are due to the observed data. Unless specified otherwise, we use uniform or flat priors for each variable on its potentially admissible range. For instance, for $\Delta g_p$ we use a U(0,2) distribution (it is constrained to be non-negative, and growth rates of 2 per week are well out of range; see Figure 2 of the main text). For parameters with no clear upper and/or lower bound we use an improper flat prior, for instance for the dispersion parameters $\phi$, which are only constrained to be positive.

The lines in Figure S1 exhibit the model inference and 95% intervals for the inferred infection rates $X_{it}$ as well as the model fit for reported COVID cases, reported COVID deaths, and total deaths ($\mu_{it}^{(\text{case/death/deathTot})}$). The dots represent the actually observed numbers ($Y_{it}^{(\text{case/death/deathTot})}$) that the model is fitted to.

There are 13 observations that have a posterior probability above 0.5 of being an outlier (all of these probabilities are in fact above 0.8 and most are very close to 1). Seven of them are zeros or negative numbers (due to ex post reporting adjustments). Six of them are otherwise extreme values; these are noted with an X instead of dot in Figure S1: two reported case rates and one reported death rate in the early days of the pandemic in Italy; two reported death rates in Spain around a time when this country had significant reporting delays and adjustments; and one reported death rate in the US state of Washington, early on in the pandemic.

Finally, we assume an AR(2) model for the progression of the idiosyncratic term in the growth rate $\Delta g_{it}^{(\text{id})}$:

$$\Delta g_{it}^{(\text{id})} = \theta_1\, \Delta g_{i,t-1}^{(\text{id})} + \theta_2\, \Delta g_{i,t-2}^{(\text{id})} + \sigma\, \epsilon_{it} \qquad \text{(S3)}$$

The AR(2) persistence parameters $\theta_k$ are included in the model based on earlier estimates of the eigenvalues $\lambda_k$ ($\theta_1 = \lambda_1 + \lambda_2$ and $\theta_2 = -\lambda_1\, \lambda_2$). We assume $\lambda_{1,2}$ = 0.9 in the main specification of the model and eigenvalues of 0.8 and 0.85 as a sensitivity check. These values are based on the range of estimated values in an earlier version of the model (for reasons of estimation speed and stability we take these values fixed in our reported estimations, ensuring that the choice of $\lambda_{1,2}$ does not materially affect our results in the sensitivity analysis reported below). The variable $\varepsilon_{it}$ follows a standard normal distribution. The standard deviation $\sigma$ is difficult to identify in the model, so it is provided as an assumption, representing the speed with which idiosyncratic changes in the growth rate tend to play out. Lower numbers make the model more sensitive to changes in policies. We assume $\sigma = 0.02$ / week, confirming in the robustness checks (Figure S2) that key outcomes do not significantly change by taking lower or higher values.

### Robustness checks

We perform several tests to check the robustness of the model. First, we calculate the autocorrelation of the residuals $\varepsilon_{it}$ of the AR(2) model for the growth rate. The lag 1 autocorrelation is around 0.1 (corresponding to an $R^2$ of around 0.01). The higher lags are even smaller in magnitude.

Second, we perform sensitivity checks on key inputs to the model: the population cut-off points for inclusion of jurisdictions; the probability of outliers $p_{\text{outlier}}$; the standard deviation $\sigma$ of the AR(2) process in Equation (S3); the exclusion of jurisdictions outside Europe and the US (*in casu* Brazil, Chile, Mexico, and South Africa); the value of the AR(2) eigenvalues $\lambda_{1,2}$; and allowing small negative values of $\Delta g_p$ (with a minimum of -0.05). For each of these changes versus the base specification, we assess the resulting impact on the policy effectiveness estimates $\Delta g_p$ as reported in Figure 2. Figure S2 displays the results. Results in red denote cases for which the base estimate is outside of the 95% posterior interval of changed specification. This situation occurs in three instances. One instance is the effect of level 2 information campaigns in jurisdictions with more than 10 million residents; apparently these campaigns might be more effective in smaller jurisdictions. The other two instances are the level 2 and level 3 testing policies (H2) when allowing negative effects $\Delta g_p$. These results suggest that these policies could be associated with a negative impact on the growth of COVID cases. Given that many of these testing policies were implemented when lockdowns were lifted, it appears likely that this negative association is due to other policy changes that were not fully captured in the OxCGRT policy database. This potential for omitted variables to affect our results is a fundamental limitation of observational studies, as discussed in the main text. Because it is plausible that the effect of increased testing is at least non-negative, we maintain that assumption in the prior for $\Delta g_p$ in our base specification.

Third, we compare actually reported COVID case numbers with out of sample predictions from the model. In order to do so, we perform an additional model run excluding the final 6 weeks of epidemiology data. Then we compare reported data of $Y_{it}^{(\text{case})}$ and $Y_{it}^{(\text{death})}$ for these 6 weeks with the posterior 95% intervals for this variable based on the model predictions excluding these data as inputs. We calculate the percentage of jurisdictions for which the actual numbers were within the 95% interval. Figure S3 shows the results. The actual data are indeed within the range of the out-of-sample predictions in close to 95% of the cases in the first weeks, with slightly lower values in some later weeks. Figure S4 shows the predicted posterior intervals for the held-out data as well as the actually reported numbers. The few instances in which the reported numbers are outside of the 95% posterior prediction range occur primarily around major policy changes. The reported numbers can be either higher (e.g. in Austria) or lower (e.g. in Belgium) than the predictions. These results are consistent with the fact that our estimates represent average effects across jurisdictions.

### Supporting Information: Tables & Figures

**Table S1.** Posterior median and 95%-interval estimates for key variables of interest, as well as Bayesian estimate statistics $n_{eff}$ and $R_{hat}$.

| | Median | 95% interval | | $n_{eff}$ | $R_{hat}$ |
|---|---|---|---|---|---|
| $w_1^{(case)}$ | 0.974 | 0.875 | 0.999 | 771 | 1.00 |
| $w_2^{(case)}$ | 0.026 | 0.001 | 0.125 | 771 | 1.00 |
| $w_1^{(death)}$ | 0.096 | 0.071 | 0.122 | 715 | 1.00 |
| $w_2^{(death)}$ | 0.438 | 0.376 | 0.508 | 430 | 1.01 |
| $w_3^{(death)}$ | 0.318 | 0.236 | 0.395 | 729 | 1.00 |
| $w_4^{(death)}$ | 0.149 | 0.086 | 0.204 | 665 | 1.00 |
| $\varphi^{(case)}$ | 6.996 | 6.137 | 7.994 | 707 | 1.00 |
| $\varphi^{(death)}$ | 7.447 | 6.285 | 8.866 | 712 | 1.00 |
| $\varphi^{(deathTot)}$ | 345.9 | 307.8 | 390.7 | 594 | 1.00 |

**Table S2**. Variance inflation factors for the policy fixed effects $\delta_{ipt}$ by policy $p$.

| | |
|---|---|
| **C1 - School closing - 2** | **3.05** |
| **C1 - School closing - 3** | 1.91 |
| **C2 - Workplace closing - 2** | 2.00 |
| **C2 - Workplace closing - 3** | 1.91 |
| **C3 - Cancel public events - 2** | 1.84 |
| **C4 - Restrictions on gatherings - 2** | 5.19 |
| **C4 - Restrictions on gatherings - 3** | 4.89 |
| **C4 - Restrictions on gatherings - 4** | 1.57 |
| **C5 - Close public transport - 1** | 1.53 |
| **C6 - Stay at home requirements - 1** | 2.78 |
| **C6 - Stay at home requirements - 2** | 2.06 |
| **C7 - Restrictions on internal movement - 1** | 3.15 |
| **C7 - Restrictions on internal movement - 2** | 1.55 |
| **C8 - International travel controls - 2** | 2.58 |
| **C8 - International travel controls - 3** | 2.79 |
| **C8 - International travel controls - 4** | 1.19 |
| **H1 - Public information campaigns - 1** | 2.16 |
| **H1 - Public information campaigns - 2** | 3.80 |
| **H2 - Testing policy - 1** | 1.79 |
| **H2 - Testing policy - 2** | 2.51 |
| **H2 - Testing policy - 3** | 1.57 |
| **H3 - Contact tracing - 1** | 2.01 |
| **H3 - Contact tracing - 2** | 1.67 |
| **C1 - School closing - 2** | 3.05 |
| **C1 - School closing - 3** | 1.91 |

**Figures**

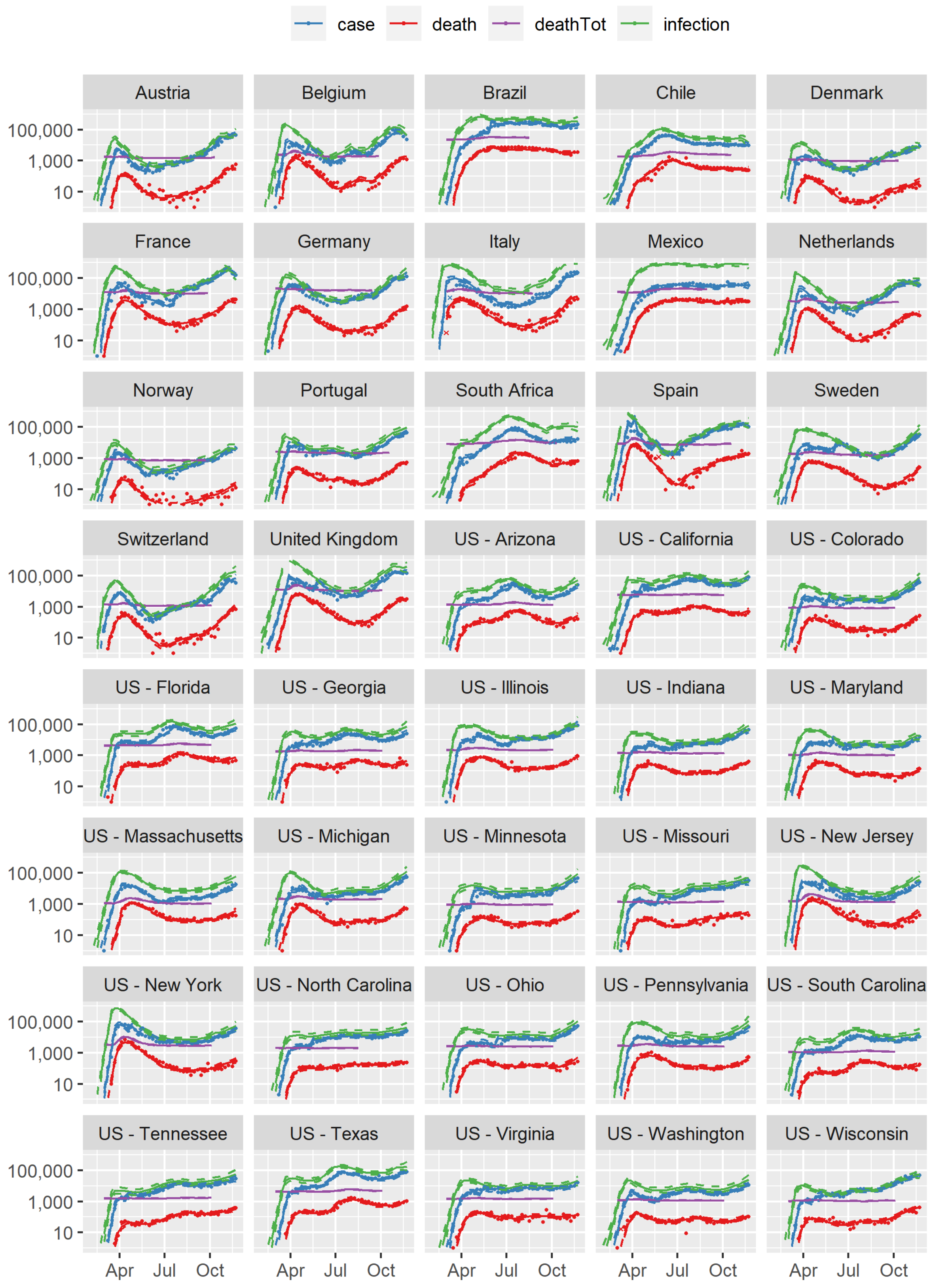


**Figure S1.** Newly identified COVID cases, COVID deaths, total deaths, and infections per week (log scale). Dots = reported; X = outlier; Solid lines = model fit; Dashed lines = 95% intervals

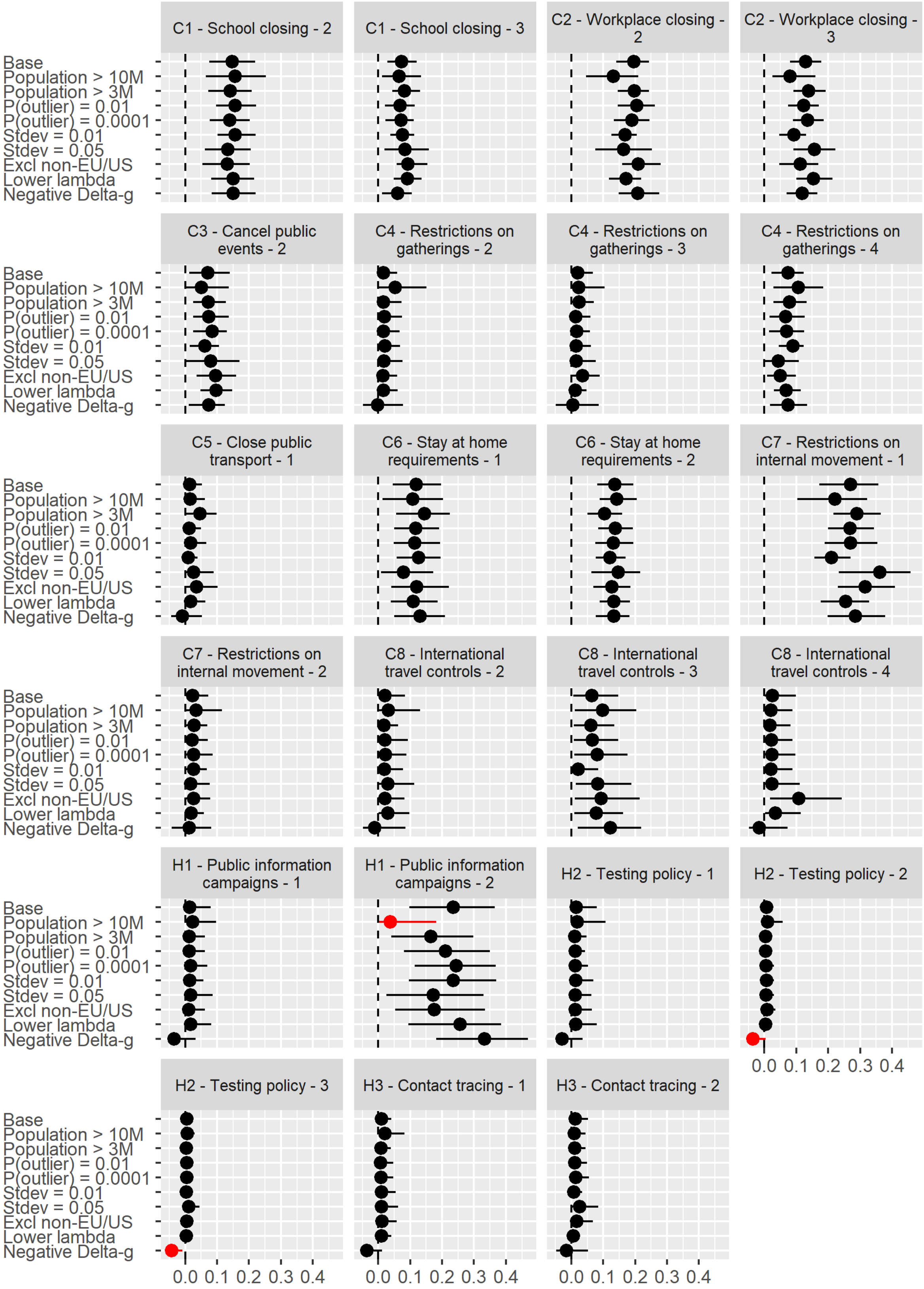


**Figure S2.** Sensitivity of policy effect estimates (as reported in Figure 2 of the main text) to different model specifications. Dots = median estimates; Lines = 95% intervals; Red = Base estimate outside of 95% posterior interval of changed specification.

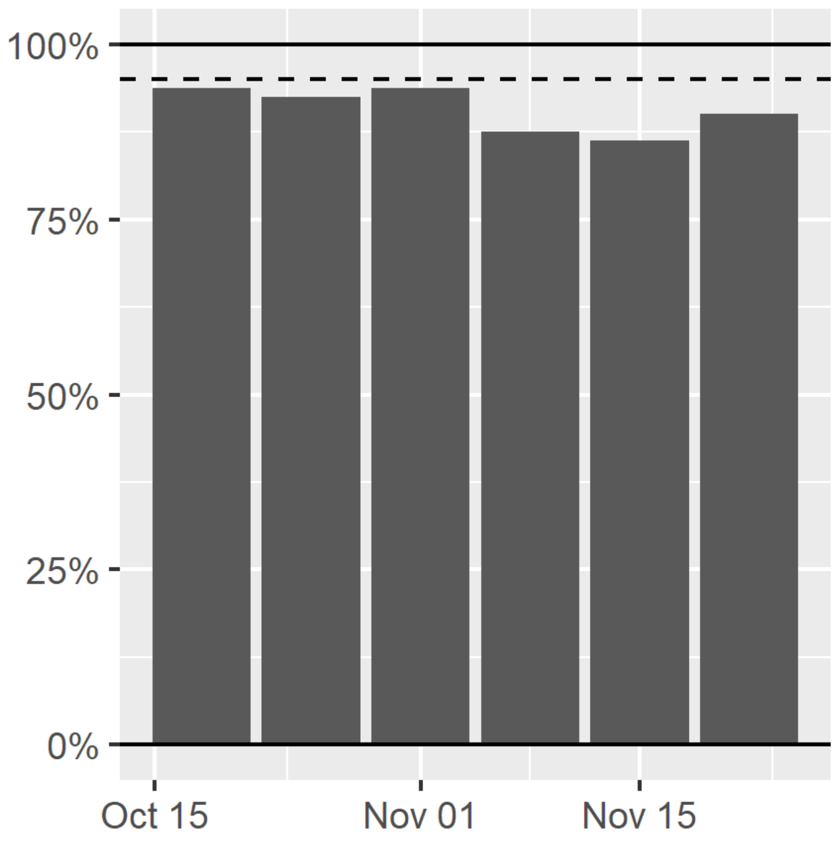


**Figure S3.** Percentage of weekly reported COVID cases and deaths within 95% interval of out-of-sample predictions from a model using input data up to 6 weeks prior to present.

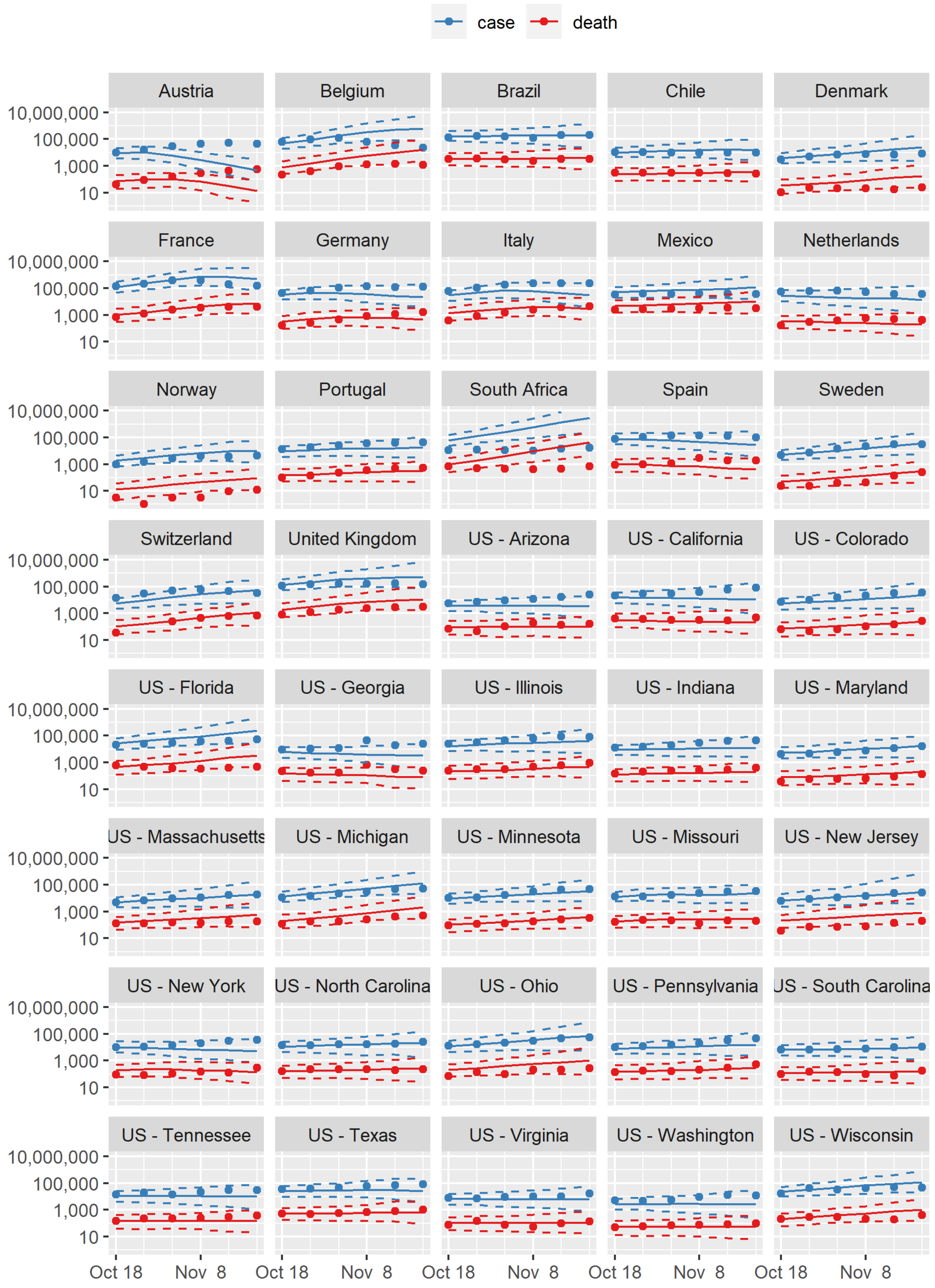


**Figure S4.** Out of sample estimates of newly identified COVID cases and deaths (log scale), with reported numbers. Dots = reported; Solid lines = model prediction; Dashed lines = 95% intervals
